# HIV Pre-exposure Prophylaxis (PrEP) Practices in Florida, USA: Clinicians’ Perceptions of Initiation, Risk Identification, Barriers, and Facilitators

**DOI:** 10.1101/2025.01.30.25321379

**Authors:** Khairul Alam Siddiqi, Shantrel S. Canidate, Yiyang Liu, Liat S. Kriegel, Sumaiya Monjur, Christa Cook, Robert L. Cook

## Abstract

This study aimed to learn clinicians’ perspectives on PrEP initiation, the HIV risk assessment process, perceived barriers to PrEP implementation, and how a potential EHR-based PrEP clinical decision support (CDS) tool can help improve their practices. Data were collected between October 2021 and November 2021 via three remote focus groups with 15 clinicians with experience prescribing PrEP. The focus groups were audio recorded, transcribed, and analyzed using thematic analysis. Five themes emerged from the qualitative analysis: (1) PrEP initiation is a joint effort between patients and clinicians; (2) Electronic health records (EHRs) are helpful but insufficient for identifying PrEP candidates; (3) Patient-clinician conversations are key for identifying PrEP candidates; (4) Patient, clinician, and system-level barriers deter PrEP implementation; and (5) Adopting technological innovations in health care can improve PrEP prescribing. Our analysis suggests that implementing effective communication strategies and behavioral interventions can improve PrEP awareness and reduce barriers in patient-clinician discussions of sexual history and substance use.

## Introduction

Pre-exposure prophylaxis (PrEP), which involves the daily use of antiretroviral medications to prevent HIV, may reduce the risk of getting HIV from sex and injection drug use by about 99% and 74%, respectively (1). A clinical practice guideline (2021 update) from the Centers for Disease Control and Prevention (CDC) provides comprehensive information for the antiretroviral PrEP, including eligibility and recommendations for medication, monitoring, and follow-up care to reduce the risk of acquiring HIV infection (2). However, PrEP awareness and uptake are low in the US, especially in the South, and among some subpopulations, including women and men who have sex with men (MSM) (3–6). Indeed, despite the availability of effective HIV prevention strategies, Florida leads the US with seven counties with high HIV burden (7), more than 115,000 people living with HIV, and around 4,500 new cases of HIV infections are diagnosed every year (8). With a significant multicultural immigrant population and tourists but restricted HIV risk and sex education and a state requirement that minors get parental consent to start PrEP, Florida poses a unique challenge to curb new HIV infections (9). Although PrEP uptake in the state has increased in recent years, disparities exist across demographics and sexual subgroups (10–12). Moreover, around 13% of people with HIV in Florida are still unaware of their HIV status, which may contribute to ongoing HIV transmission. Thus, there is a need to implement evidence-based HIV prevention interventions at healthcare facilities to strengthen PrEP practices in states with high HIV incidence, like Florida.

A complex array of structural, social, clinical, and behavioral barriers at the patient and clinician level contribute to the lack of PrEP uptake (13–15). Addressing these patient-level barriers — including perceived HIV risk, stigma, low PrEP awareness, concerns about side effects and effectiveness, and cost of PrEP — that affect access and adherence to PrEP and other HIV prevention strategies is one key to increasing PrEP usage (16). In addition, several clinician-related barriers have been identified, and these could vary in different practice settings. Clinicians have limited time to collect diverse patient information or may feel uncomfortable asking about sexual history during healthcare visits (17–19). Thus, providing educational materials, training on PrEP prescribing, and appropriate clinical resources for clinicians and creating a judgment-free environment for patients to talk about sensitive issues (e.g., sexual history and substance use) can improve PrEP practices (15).

Electronic health records (EHRs) include rich patient health data that clinicians can use to identify individuals at increased risk for HIV, including potential candidates for PrEP. An EHR-based risk prediction tool that assesses demographic, clinical, and non-clinical characteristics has the potential to automatically identify people with a high risk of HIV acquisition who may benefit from PrEP. Studies have shown that clinicians with the best clinical decision support (CDS) tools can improve care quality and utilization because such tools provide knowledge ranging from up-to-date clinical guidelines to patient-specific information, enabling them to make smarter, quicker, and more informed decisions (20–22). However, various user (e.g., lack of knowledge, training, time) and system level (e.g., workload, hard technology, lack of hardware, data) barriers could retard the implementation of CDS for PrEP in clinical practices (23). Recent studies demonstrated that addressing these drawbacks and engaging users in the development and deployment of an EHR-based PrEP CDS tool could facilitate patient-provider communication about HIV risk, help patients accurately perceive their risk, destigmatize and standardize risk assessment, and prospectively identify potential PrEP candidates, which can improve PrEP counseling and initiation (24–27). Very little of this recent evidence was available at the time we planned and conducted the study, and few of these studies focused on identifying gaps in current practices that should be considered during the development and implementation of a CDS. Therefore, our study highlighted clinicians’ perspectives on PrEP initiation, the HIV risk assessment process, perceived barriers to PrEP implementation, and how a CDS tool can help improve their practices. Learning insights from the experience of primary care and specialty doctors, nurses, and pharmacists working in unique care settings regarding patients from diverse cultures and huge older and tourist populations would be valuable.

## Materials and Methods

### Study Participants and Setting

A purposeful sample of clinicians across various specialties and clinical settings with experience prescribing PrEP in Florida were identified and recruited to participate in three virtual focus groups. One hundred and ten clinicians were invited to participate in the study through the email lists of two professional research networks: OneFlorida+ Clinical Research Network and the Southern HIV and Alcohol Research Consortium. The first fifteen clinicians (11 doctors, 2 nurses, 2 pharmacists) who consented to participate were recruited and subsequently assigned to one of three focus groups based on their availability or convenience (five participants per focus group). Nurses and pharmacists were included because they have key roles in prescribing and distributing PrEP under collaborative practice agreements in Florida. With three focus groups conducted, we observed repetitive discussion around the topics from the participants; thus, we believe saturation was reached. Before each focus group, participants completed an online demographic survey (e.g., age group, specialty, years of experience) and signed an informed consent form that explained the study purpose, focus group topics, the risks and benefits of participation, confidentiality, and compensation.

### Focus Group Procedures

The focus groups occurred between October 2021 and November 2021. Each discussion lasted about sixty minutes and was conducted on the Zoom platform using a semi-structured discussion guide. All participants were given a code name (e.g., blue, gold, circle) before joining the Zoom meeting. The participants also had the option to keep their cameras off to maintain anonymity. A moderator, a note-taker, and a principal investigator also attended each focus group. The moderator provided the focus group with general etiquette guidelines in the Zoom environment. As an icebreaker, participants were asked to talk briefly about their work environment. Data collection questions included, How do you typically identify patients in need of PrEP services? What do you think are the barriers to prescribing PrEP? What are your thoughts about using an EHR-based CDS tool to assist with providing PrEP services? To minimize the effects of any group participation bias, the moderator tried to engage all participants in the discussion. After completing the focus group, each participant received a $50 e-gift card.

### Ethics Statement

This study was reviewed and approved by the Institutional Review Board of the University of Florida (#IRB202101059), and the procedures followed were per the Helsinki Declaration as revised in 2013. The participants provided written informed consent to participate in this study.

### Data Analysis

The focus groups were audio recorded, transcribed verbatim, checked for errors, and analyzed using inductive thematic analysis involving deriving meaning and identifying themes from data without preconceptions (28). A consensual coding process wherein two coders independently coded the transcripts using NVivo was utilized. The two coders first generated and defined an initial set of codes as they emerged from the initial transcript. Initial codes appeared iteratively during the line-by-line review of the transcripts and field notes. The coding team grouped together codes that represented similar concepts and themes and developed an initial coding scheme. The coding team met regularly to discuss and refine the coding scheme, including adding or deleting codes, updating code descriptions, and identifying quotes that best represented the specific codes. Considering the small sample of data and different numbers of codes in different topics of interest (29), we calculated the percentage of agreement between two independent coders to make sure it reached a substantial agreement, i.e., at least 80% across topics (30). The codes that emerged from the transcripts were compared, discussed, and refined by resolving discrepancies to finalize the codes. The codes were eventually organized to reflect similar concepts or patterns in order to develop themes that answered our study questions.

## Results

### Participant characteristics

Of the 15 clinicians who participated in the three focus groups, 73% were medical doctors (*n*=11), 60% were male (*n*=9), 47% were White (*n*=7), and 33% were 30-39 years old (*n*=5). Most of the clinicians worked in primary care (n=13), and their average work experience was about ten years (**Table 1**). In the focus group discussions, only two clinicians mentioned that they independently initiated PrEP prescribing for the first time without patient prompting, while other clinicians mentioned that they mainly initiated PrEP when the patient asked for it (*n*=5). Four clinicians reported that they initiated PrEP prescribing as part of their job responsibilities, and they received training before it. Seven clinicians indicated that they use the PrEP prescribing guidelines from the Centers for Disease Control and Prevention (CDC) in their practices, while few others talked about other checklists or questionnaires that they follow to identify HIV risks and PrEP candidates. Five major themes emerged during the analysis of the transcripts reflecting the latest PrEP prescribing practices in Florida, including the PrEP initiation process, the use of EHRs in HIV risk assessment, patient-clinician interactions to discuss risk factors, major barriers to PrEP implementation, and adopting technological innovations to overcome those barriers. The key subthemes discussed under these major themes are illustrated in **Table 2** and in detail in the following paragraphs.

**Table 1.** Demographic characteristics of the clinicians (n=15)

| **F**<br>**G**<br>**#** | **Codena**<br>**me** | **Specialty** | **Race/Eth**<br>**nicity** | **Gender** | **Age**<br>**group** | **Setting** | **Years**<br>**prescribed**<br>**PrEP** |
| --- | --- | --- | --- | --- | --- | --- | --- |
| 1 | Green | Family medicine<br>doctor | White | Female | 30-39 | Primary<br>Care | 7 |
| 1 | Orange | Advanced<br>practice<br>registered nurses | White | Female | 50-59 | Primary<br>Care | 2 |
| 1 | Purple | Internal medicine<br>doctor | Asian | Female | 40-49 | Primary<br>Care | 10 |
| 1 | Yellow | Family medicine<br>doctor | White | Male | 30-39 | Primary<br>Care | 2 |
| 1 | Red | Family medicine<br>doctor | White | Male | 50-59 | Primary<br>Care | 2 |
| 2 | Diamond | Pharmacist | African<br>American | Male | 20-29 | Primary<br>Care | 2 |
| 2 | Triangle | Pharmacist | African<br>American | Male | 30-39 | Primary<br>Care | 2 |
| 2 | Circle | Infectious disease<br>expert | White | Male | 50-59 | Infectious<br>disease<br>clinic | 4 |
| 2 | Square | Adolescent<br>medicine expert | White | Female | 30-39 | Pediatric<br>hospital | 3 |
| 2 | Star | Internal medicine<br>doctor | Asian | Male | 20-29 | Teaching<br>hospital | 2 |
| 3 | Grey | Family medicine<br>doctor | White | Male | 50-59 | Primary<br>Care | 5 |
| 3 | Blue | Internal medicine<br>doctor | Asian | Female | 40-49 | Primary<br>Care | 5 |
| 3 | Silver | Internal medicine<br>doctor | African<br>American | Male | 20-29 | Teaching<br>hospital | 2 |
| 3 | Mint | Internal medicine<br>doctor | White<br>(Hispanic<br>) | Female | 30-39 | Primary<br>Care | 4 |
| 3 | Gold | Advanced<br>practice<br>registered nurses | African<br>American | Male | 20-29 | Urgent care | 2 |

**Table 2.** Overview of key themes and sub-themes.

|  | **Theme** | **Sub-theme** |
| --- | --- | --- |
| 1 | PrEP initiation is a joint effort between patients and clinicians | Patients take a preventive approach when aware of risk and PrEP<br>Clinicians initiate PrEP discussions when patients indicate HIV risks<br>CDC PrEP guidelines are helpful, but they need a comprehensive screening tool |
| 2 | EHRs are helpful but insufficient for identifying PrEP candidates | EHRs can pool HIV risk factors from previous lab results<br>EHRs can help choose the right medication types and dosage<br>EHRs can be used to collect sexual risk behavior data<br>Use of EHRs is limited in PrEP practices |
| 3 | Patient-clinician conversations are key for identifying PrEP candidates | Talk between patient and clinician helps reveal HIV risks<br>Male patients are more open to HIV-related discussion<br>Stigmas affect quality discussion between patients and clinicians |
| 4 | Patient, clinician, and system-level barriers deter PrEP implementation | Follow-up to PrEP continuation is challenging<br>Perceived low risks among patients and clinicians are key barriers<br>Lack of HIV data in EHRs interrupts risk screening<br>PrEP reimbursement is difficult for some clinicians |
| 5 | Adopting technological innovations in health care can improve PrEP prescribing | Telemedicine can increase patient engagement<br>An automated alert or reminder tool can help manage PrEP prescribing and follow-up<br>CDS tool should be a specialty and setting-specific<br>Patient and clinician's perspectives should be incorporated into the CDS tool |

## 1. PrEP initiation is a joint effort between patients and clinicians

The clinicians indicated that PrEP initiation could be led by both them and their patients. They consider initiating PrEP discussion when patients indicate any HIV risk factors, including sexually transmitted infections (STIs), unsafe sexual practices, and substance use. Patients often approach their doctors as an HIV prevention resource when they perceive themselves to be high risk and are aware of PrEP through advertisements or internet searches (e.g., radio, TV, billboards, and social media).

“Sometimes they’re (patients) more knowledgeable, maybe…they read about it before coming, and …sometimes they’re more at risk. Maybe there’s a bigger population where they’re more at risk, like… if they have multiple partners, MSM, or – like they know they should come more frequently, and these persons, sometimes they’re knowledgeable about a PrEP, and …they would come for that. It is the main complaint, like this is why they’re coming today, to discuss it.” (Doctor, female, white, and 30–39 years old)

Some clinicians received PrEP training and education to accommodate client-led requests. Training makes clinicians aware of identifying risk factors among patients and linking them to HIV prevention care.

“So, most patients are self-selected,…but also training people (clinicians) to be able to among the census of their population they’re taking care of to bring it up to people at their routine visit if they look like (they are at-risk), they would be a candidate for PrEP.” (Doctor, male, white, 50–59 years old)

While proactive attempts by patients to initiate PrEP are part of clinical practice, clinicians are also well-situated to identify the right candidates and to initiate PrEP discussions.

“When a patient comes to the hospital, either he or she has STD, I can be able to actually initiate a conversation about the sexual practices and what they have been actually – how many partners they’ve been engaging with. So definitely, that is the main method that we – I actually, me personally, I start the conversation about PrEP. I try to educate the person about PrEP.” (Doctor, male, African American, 20–29 years old)

However, focus groups revealed that some clinicians do not follow a standardized procedure to collect patients’ risk information, so their PrEP conversations could be improved if they were supplemented with a standard sexual health protocol. Some clinicians use tools, like self-made sexual history forms, to record patients’ HIV risks, such as having multiple partners with no constant condom use, engaging in sexual activity while on alcohol or drugs, and the presence of multiple STIs. These histories are also accompanied by a note template or questionnaire, adopted in the EHR or separate from it, to ensure all relevant information is collected.

“I would choose something maybe that I’ve made. Like, if I want to know a history of past STIs, if this treatment has been done before, or has been used before, if they were on PEP (post-exposure prophylaxis), or PrEP, or like some information, we want to have it, and we don’t want to miss any of that. So, maybe I would use a prepopulated, or like a pre-prepared questionnaire, or a set, and fill it, like during the encounter.” (Doctor, female, white, 30– 39 years old) Standard clinical practice is often informed by the CDC guidelines (Center for Disease Control and Prevention, 2021) that emphasize assessing patients’ sexual and substance use behaviors to identify the risk of HIV acquisition; however, not all clinicians agree that these guidelines are sufficient for the identification of HIV risk and PrEP need.

“…I’ll talk about screening – you know, the CDC guidelines for screening for gonorrhea, chlamydia, for HIV, for hepatitis C, and then I talk about risk factors, and that’s when – the time we identify potential for PrEP,…” (Doctor, female, Asian, 40–49 years old)

Following the CDC’s guidelines is not easy for some clinicians because it looks tough at first glance—they find it especially daunting to read a hundred-page recommendation amidst their busy schedules.

“Yeah, the CDC guidelines were quite intimidating with primary care doctors. I mean, it’s crazy… primary care doctors are scared to death to do this until they realize how easy it is to do it and looking at the CDC guidelines really wouldn’t actually make that point.” (Doctor, male, white, 50–59 years old)

## 2. EHRs are helpful but insufficient for identifying PrEP candidates

Some of the clinicians found EHRs to be very effective for identifying PrEP candidates because they can screen HIV risk factors through patients’ current and previous lab test results and diagnoses during healthcare visits.

“I’m using the medical record to look back and see if they’ve had the right labs or if they’ve tested positive for STIs in the past, then that’s probably how I use it.” (Doctor, female, white, 30–39 years old)

EHRs can also be useful for directing clinicians toward the right PrEP medication and dosage based on the latest policy guidelines. One clinician commented on this:

“…I have to remember ‘Okay, which one is approved for male? And which one’s approved for female?’ So, it’s something that the EMR (electronic medical record) could do a better job of guiding my decision making, but I’m going to stick with the approval and the guidelines.” (Doctor, male, white, 50–59 years old)

Clinicians often seek out histories of STI screening, PEP or PrEP use, and substance use in the EHRs as these may indicate patients’ past or present risks to HIV acquisition.

“…the way I would use a computer is to see myself how many times the patients had STIs and how many times the patient has had asked for an HIV test or asked for STI screening.” (Doctor, male, white, 50–59 years old)

Likewise, EHRs that include patient’s sexual history (e.g., number of partners, condom use) and gender identity can provide important insights when identifying PrEP candidates. According to the clinicians, HIV risks and risk perceptions vary with sexual orientation and gender identity among both patients and clinicians.

“In the EMR at our center, we have a tab, or a heading, … and that’s the historical page where we have sexual history and preference and partners. We actually just recently upgraded it in the last year or two; we have sexual orientation, gender identity on a separate tab, and these things are useful.” (Doctor, male, white, 50–59 years old)

While some clinicians think EHRs are useful in HIV risk screening and PrEP prescribing, some of the clinicians, felt that EHRs are insufficient, given the limited records available due to privacy restrictions and inadequately collected medical records.

“So, even like if someone else has seen the patient and they’re following some confidentiality, privacy rules, we’re not going to be able to reach these – this information” (Doctor, female, white, 30–39 years old)

Furthermore, some clinicians emphasized the lack of HIV risk behavior data in the EHRs. In fact, when clinicians can access patient records, they are sometimes limited by what data is actually available. One clinician indicated:

“And even in the EMR world, we’ve been on the EMR for about a decade, a lot of the past records don’t really have the details we want related to risk factors, because there’s no details about sexual history, social history. Those don’t get included in past records, and if they are, get buried in 600 pages of past records.” (Doctor, female, Asian, 40–49 years old)

## 3. Patient-clinician conversations are key for identifying PrEP candidates

In addition to the patient records available in the EHR, clinicians may ask their patients about their partner’s HIV status, potential other contacts in case they are HIV positive, number of partners, age at first intercourse, condom use, and whether the patient’s partner is on an antiretroviral drug. One clinician commented that this information is not well documented in EHRs:

“Just in general, sexual practices that aren’t asked or definitely not documented well enough, but like number of partners, age at first intercourse, something I think about a lot for risk factors, condom use.” (Doctor, female, white, 30–39 years old)

To describe a potential PrEP candidate, one clinician spoke about how they approach a serodiscordant couple to initiate PrEP discussion.

“So, a patient I saw a couple of weeks ago came in with his partner who was – they were a serodiscordant couple and the partner was not on PrEP… And so there was just an easy, perfect opportunity without having to go behind anybody’s back because they’re both sitting there.” (Doctor, male, white, 50–59 years old)

In addition to talking about sexual health, clinicians also initiate PrEP discussions if their patients indicate substance use or needle sharing as part of standard practice. These indications are either highlighted through a medical history review or during conversations with their patients.

“I also have a lot of patients who I treat with substance use concerns and so, for a lot of them who are maybe using or sharing needles or just engaging in risky sexual practices, often who have had multiple STIs,…” (Doctor, female, white, 30–39 years old)

The clinicians further shared their perceptions regarding gender differences that influence initiating PrEP discussion. In most cases, they perceive that men who have sex with multiple partners or who engage in anal sex are at higher risk for HIV infection. Consequently, they approach their male patients more often regarding PrEP. They also find that men are generally more receptive to HIV-related discussions than women, resulting in greater confidence in communicating with male patients.

“…I just find, at least historically, at least with my patients, male patients either are more receptive to that – opening up that conversation or considering it than the female patients that I’ve had.” (Doctor, female, white, 30–39 years old)

Another clinician echoed a similar experience, indicating that males are more confident about their situation.

“I’m not sure if maybe it’s sexuality preferences and the kind of behavior that they’re engaging with, I feel like some of the really open patients that I’ve had are very candid and very open. They’re almost uninhibited in terms of discussing what kind of behaviors they engage in versus a lot of the female patients that I think merited that conversation. They kind of shut down very quickly. So, I don’t know if it’s embarrassment. I don’t know if it’s, no, no, no, like maybe a little bit of self-denial. I’m not really sure, but that’s always the experiences I’ve tended to have.” (Nurse, male, African American, 20–29 years old)

These perceptions are also influenced by social stigmas related to sexual history and drug use, resulting in a general reluctance of some clinicians to ask questions related to patients’ sexual practices.

“So, maybe if you see someone’s coming in for concern for infection, STD infection and you’ve seen – that’s all they come in for, that needs to have a more thorough discussion in terms of their sexual activity and potential exposure risks. Sometimes, they’d like to gloss over kinds of sensitive topics of conversation. So, sometimes we’ll see patients who are actually that is a concern, but no one ever asked them about it. Nobody asked them if they were sexually active. Nobody had asked them if they engaged in sexual activity with male or female partners or multiple partners. And they were too embarrassed” (Doctor, female, white, 30–39 years old)

## 4. Patient, clinician, and system-level barriers deter PrEP implementation

The requirement of a follow-up visit was identified as a prime barrier to initiating as well as continuing PrEP because patients consider managing a follow-up visit and going through related sexual health screening every three months inconvenient. The clinicians noted that even if they prescribe PrEP, successful implementation requires buy-in from their patients, given that PrEP requires continued use throughout risk exposure. Patients are also sometimes deterred by the potential side effects of the PrEP medications. One clinician shared how these inconveniences influence patients’ decisions to take PrEP:

“…say you start them on PrEP and you’re supposed to monitor them afterwards, because this is a barrier I’ve found is like, some patients that maybe were initially interested when they hear like, oh, I have to come back every three months and I’m supposed to get checked for HIV and STDs again, they’re like, I’m not that interested anymore. And especially women if they think that it’s going to be necessarily repeat pelvic swabs every three months, like, they may not be so interested. I could see that being a barrier.” (Doctor, female, white, 30–39 years old)

Another clinician extended the discussion with observations regarding the lack of adherence to and side effects of PrEP:

“So, … one of the biggest barrier is like follow-up adherence, compliance to medication and then safety monitoring, … but so, those factor in quite a bit and I think that’s where it, charts can also be helpful in trying to assess especially if they already have a predisposing bone or kidney problem or something that would require some frequent monitoring.” (Doctor, male, Asian, 20–29 years old)

There is also a broader lack of knowledge and awareness about PrEP among patients and clinicians. Some patients do not consider themselves to be at risk and some clinicians are indifferent in prescribing PrEP. Moreover, some clinicians perceive that it’s not their responsibilities, especially those from non-HIV specialties or clinics.

“…I’d say the thing that from what I’ve seen in training docs (doctors) is that they, a lot of them don’t realize that they can do it. And I think that it’s more of an issue for them to realize you don’t have to be an HIV treating physician in order to prescribe PrEP that’s probably the biggest thing I’ve seen.” (Doctor, male, white, 50–59 years old)

The clinicians further contended that finding the right PrEP candidate is challenging when patients perceive themselves as not at risk, HIV risk information is not available in the EHRs, and they lack an appropriate questionnaire to collect sexual history. Some clinicians may also lack an HIV risk assessment tool in their practices.

“Definitely, I’ll say not having a questionnaire or something that actually can be able to make you collect exact sexual history information about a particular patient, not having that actually makes it hard for a clinician to be able to actually identify a person that really needs the PrEP.” (Nurse, male, African American, 20–29 years old)

In addition, some of the clinicians noted that accessing patients’ previous medical records from other healthcare systems, managing them, and finding desirable data are very challenging.

“So, I routinely don’t ask for previous records because the most egregious example is the patient came to me from a Department of Defense Hospital, and the previous records totaled over 600 pages, and it was $15 of postage alone just to send it to us, and the dilution of relevant data was so profound. I don’t know what we frankly did with the papers, but we stopped asking for old records pretty quickly after that.” (Doctor, male, white, 50–59 years old)

Finally, the clinicians cited funding as a critical barrier to PrEP implementation. Although PrEP is free or low-cost for individuals with health insurance and through some PrEP assistance programs, getting reimbursed for PrEP is often difficult for clinicians, indicating numerous paperwork related to justifying a bill for PrEP.

“I don’t know what you guys’ experiences are, but I have difficulty getting funding for PrEP. It is a lot of administrative burden on my end at least, from the insurance side, to get on PrEP. Because – a lot of documentation from the insurance companies, or prior authorizations, or peer-to-peer. So, I think that’s a huge barrier.” (Doctor, female, Asian, 40–49 years old)

## 5. Adopting technological innovations in health care can improve PrEP prescribing

While the clinicians discussed several barriers to PrEP implementation, they also offered potential solutions to improve PrEP practices in Florida. To address broad stigmas and access to sexual health care, the clinicians suggested that telemedicine, computer-based CDS tools, and community-based approaches could potentially increase patient engagement.

“Yeah, I mean, from the patient perspective, access is – these is – this is a population that is very difficult to bring them back, and we should make access either with telemedicine, do telephone calls. So, it should be billable, right? We should be able to do that instead of having them come in person, go to their workplace, go do their location in the community. Those are – I think would remove some barriers to access.” (Doctor, female, Asian, 40–49 years old)

They think that these technological innovations can help resolve some barriers and create a safe and judgment-free environment to talk about HIV risk behavior, including sexual history and substance use:

“If we could have an open place where it’s virtual like here. And we do like a session like this without the video being open. And maybe just using the voice over, people will be open, and maybe we could get to have more because I think they’re afraid of being judged. Maybe if there’s a place where you can’t see them, where they cannot be vulnerable to you, that will help in giving out more of the PrEP.” (Doctor, male, African American, 20–29 years old)

Another clinician indicated the need for automated tracking of patients’ risk exposure and a reminder to update risk information over time, which may help clinicians manage patient follow-ups.

“…But if there was some way to automate the reminder that says once we prescribe PrEP, there’s sort of this quarterly obligation to – or, at least, best practice, to check the labs, and reassess their STI status quarterly, what are the things we can do to automate that process when I don’t have their chart open? Because that’s a potential lapse in care, and that’s easy for computers to do, that’s really hard for providers to do. So how do we automate that?” (Doctor, male, white, 50–59 years old)

Furthermore, the clinicians thought that an automated CDS tool could play a critical role in facilitating PrEP prescribing practices. Indeed, based on positive experiences with other CDS tools or systems for managing other conditions (e.g., cardiovascular disease [ASCVD calculator], vaccinations, well-child visits and avoiding duplicate laboratory tests [BPA-Best Practice Advisory]), several clinicians recommended developing a CDS tool to manage PrEP prescriptions. “Yeah… ASCVD calculator, which calculates a patient’s cardiovascular disease score and even give us a percentage value with their risk factors above and that really guides our management for prescription of statin therapy. It’s super convenient. So, now, I feel like a tool in this realm may also be helpful for diagnosing PrEP.” (Doctor, male, Asian, 20–29 years old)

Since CDS tools put all required information together in one place, they save clinicians a lot of time and effort compared to manual EHR review or paper-based risk assessments and help them focus on clinical decision-making.

“So, I’d say it’s kind of – it saves lot of time compared to the older method where we used to use files and everything. It helps predict patterns in certain patients, especially patients with lifetime diseases, such as diabetes and such. So, I’d say they’re very effective, yeah, in predictable patterns.” (Doctor, male, African American, 20–29 years old)

However, a CDS tool can be inappropriate when it is not customized for the clinical practice and is adopted outside of the EHR system because it may increase unnecessary workload. Furthermore, when the CDS tool is not disease- and visit-specific, it can become irrelevant for some practices. Thus, the clinicians expressed mixed feelings about using an existing CDS tool in their practices: “So, I work in two different systems, one that has an EHR that uses the clinical decision tools, or what we call BPAs, very strategically, and then I’m in the other EHR system, which the BPAs are way too much. So, I do – I have a love-hate relationship to them because of my experiences. I will tell you, I think the ones that are really helpful in terms of preventive screening are not as great, because we live it, and we breathe it, so we don’t need those. But the ones that are not as well known, maybe like PrEP, or opioid overuse risk, those may be more helpful. But ones that keep me up for health maintenance is very painful. So, I think there’s a delicate balance here.” (Doctor, female, Asian, 40–49 years old)

Therefore, they suggested designing a PrEP CDS tool that is based on the one relevant, successful EHR-based CDS so that it is familiar to clinicians. They also recommended customizing the PrEP CDS tool for different practice areas (some clinicians do not prescribe PrEP) and service needs (all visits may not need PrEP).

“So, I think that it would be potentially very harmful to create like a BPA that just pops up on any random visit, for any random provider… You know, getting that alert every single time would probably almost discourage them even more from taking care of these vulnerable populations. So, I think that’s something you have to be careful and cognizant about”. (Doctor, male, white, 30–39 years old)

In addition, the clinicians emphasized ensuring the reliability of the information that the tool will use, protecting patient privacy, making it user-friendly, and customizing it for certain practices:

“Yes, yes, of course. To me, I think it will be very useful as long as the privacy of our patients is protected and we have, because you understand that there are a lot of stigmatizations to people using PrEP to some extent and well, if we try our best to ensure that the tool is more private, and the anonymity of our patients is kept.” (Pharmacist, male, African American, 30–39 years old)

Finally, they thought that the development of a PrEP CDS tool should incorporate clinicians’ concerns regarding data quality, potential data breaches, increased workload, and potential challenges associated with integrating sexual history data:

“I think that would be reasonable, and if we had good data, granular data that we could pull out of the EMR, I think that population-level intervention makes sense.” (Doctor, male, white, 50–59 years old)

## Discussion

Given the consistently higher rates of new HIV infections and lower rates of PrEP use in Florida, our study provides useful information about the gaps in current PrEP practices and potential solutions for PrEP promotion. Our study highlighted that patient-provider collaboration, the use of EHRs, the importance of risk communication, and technological innovations are pivotal to improving HIV risk screening, identifying potential PrEP candidates, and initiating PrEP during healthcare visits. During the time we collected data, there were a few other similar studies, and some that have been published in recent years mostly focused on digital innovations in promoting PrEP (24–27), while our study focused on identifying strengths and gaps in current practices and discussed digital innovations as a potential solution to improve PrEP prescribing.

Our analysis showed the active role of patients in initiating PrEP discussions alongside clinician-led regular PrEP practices. Indeed, the clinicians participating in our focus groups indicated that when patients are aware of their risk of acquiring HIV and know about PrEP from social media, websites, or any other sources, they proactively initiate PrEP discussions with their healthcare providers. The role of mass media in creating awareness and increasing the uptake of PrEP is well documented (31, 32). Our clinicians also acknowledged that their first PrEP initiations were led by their patients’ intentions to have a PrEP prescription. However, when patients are not aware of their risks, clinicians can play a critical role in assessing their risks and initiating PrEP discussions. These findings indicate the importance of shared decision-making in promoting PrEP, which may help to reduce stigma related to HIV/PrEP, promote cultural competence and humility, build trust, and maximize patient autonomy (33, 34). However, given that individuals belonging to marginalized populations could be uninformed and less comfortable initiating discussions about risk behaviors, clinicians should prompt the preventive discussion by adopting status-neutral HIV care (35).

Our study also identified that clinicians use EHRs to find their patients’ current and previous lab test results and diagnoses indicative of HIV risks, including records of STI diagnoses and treatment, PEP and/or PrEP use, and substance use. However, although the Health Insurance Portability and Accountability Act (HIPAA) regulations allow providers to share EHRs with another provider with their patient’s authorization (36), our clinicians found that accessing previous medical records from other healthcare systems was challenging. Additionally, they indicated that retrieving HIV risk factors from numerous records is difficult. However, CDS tools have the ability to analyze large volumes of structured and/or unstructured data within minutes and can predict the likelihood of HIV acquisition (37–39). The literature shows that CDS tools are increasingly being used in HIV prevention and care and that they are considered successful HIV prevention interventions (26, 40, 41). Accordingly, our analysis found that the clinicians that we met with were enthusiastic about a future PrEP CDS tool, likely because of positive experiences with existing CDS tools that save time by summarizing patient information and predicting certain conditions. Based on their experiences, they further suggested modeling a PrEP CDS tool after an effective existing tool and customizing it to accommodate different specialties, practice types, and visits. However, our clinicians also voiced concerns about data quality, potential data breaches, collecting sexual history data, integrating data from multiple sources, and the increased workload to run a new tool. While it is critical to overcome these data issues, incorporating patients’ perspectives in developing and adopting such a digital PrEP tool is the key to successful implementation into clinical practices (42, 43).

Several roadblocks to collecting appropriate data and identifying individuals who may benefit from PrEP, including clinician- and patient-level stigma, lack of awareness, and lack of data sharing across healthcare settings, were also discovered during our analysis. To overcome this challenge, some clinicians use pre-populated checklists or questionnaires to collect information about sexual history, STIs, and substance use, which may indicate risks of HIV acquisition. Clinicians also mentioned that they follow the CDC guidelines and collect information about sexual history and other PrEP prescribing criteria using a template or from EHRs. Clinical practice guidelines from the CDC reflect the latest evidence-based practices and US Food and Drug Administration (FDA) recommendations regarding identifying patients who are eligible for PrEP and the correct drug regimens (2). Our analysis suggests that many clinicians found the CDC’s PrEP clinical practice guidelines helpful during their practices, while others expressed struggles with them.

While regular follow-ups improve patient outcomes, our study identified that loss to follow-up is a major barrier to successful PrEP implementation. Indeed, having a mandatory follow-up checkup and related sexual health screening every three months may discourage patients from continuing PrEP use. Previous studies have identified that regular follow-up visits and associated testing are challenging for patients (44–46). Our study suggested that telemedicine or an automated CDS tool may help engage patients with more care by removing the discomfort associated with discussing their sexual history during visits. It has actually been shown that patients prefer follow-up care by phone or email over in-person visits and think a reminder can help them stay on track (47). Moreover, since PrEP continuation and follow-up visits are associated with higher costs of care, having healthcare coverage is critical. Consistent with previous studies (48–50), our study identified concern about cost as another possible barrier to PrEP implementation.

Although clinicians reported that some patients are not comfortable talking to clinicians about their sexual history, some clinicians proactively initiate conversations with patients about their partners’ HIV status and potential other risk behaviors. Nonjudgmental discussions between patients and clinicians are critical for initiating conversation, collecting accurate risk information, and encouraging patients to avoid HIV risk behaviors (34, 51). Yet, sometimes clinicians lack appropriate clinical resources, including questionnaires or checklists, to lead these discussions in the right direction and prescribe PrEP (15). Moreover, both our male and female clinicians reported stigma surrounding discussions of sexual history or drug use with their patients, especially their female patients, resulting in incomplete data and improper risk assessments. Indeed, the perception that female patients are less receptive to PrEP conversations could be a potential barrier to promoting PrEP among women. Several studies demonstrated clinicians were less likely to prescribe PrEP for women (6, 52, 53). Furthermore, clinician-level stigma or indifference to sexual history taking can exaggerate the masking of patients’ HIV risks (54, 55). Therefore, a standard survey instrument for patients to collect risk information and a CDS tool for clinicians to prescribe PrEP can improve the current workflow without interfering with their privacy and comfort zones.

Our study has several limitations. First, the sampling of participants was purposive, and representation favored one healthcare system. Thus, our study findings may not be generalizable to all healthcare settings in Florida or elsewhere. However, representativeness was not the goal of our sampling, and we prioritized interviewing clinicians with experience in prescribing PrEP.

Second, clinical contextual elements, such as the specialty of the clinicians and clinic settings, were outside the scope of this study. Given the importance of these elements in determining PrEP practices and EHR usage, our future study considered these to develop and tailor a PrEP promotion intervention. Next, the focus groups were held virtually, and three participants turned their cameras off during the discussions (turning the camera on was not mandatory), which may have impacted the quality of the discussions. Furthermore, due to meeting virtually, some aspects of the data collection, such as body language, may not have been captured. Overall, we observed that the participants were more involved in the discussions when their cameras were turned on.

## Conclusions

Our analysis suggests that using additional tools alongside the CDC guidelines to assess patients’ HIV risks and identify the right candidates for PrEP may help clinicians in their practices. Furthermore, employing effective communication strategies and behavioral interventions can improve the health literacy of both patients and clinicians and also promote productive patient-clinician discussions of sexual history and substance use. There should also be smoother data-sharing processes between different providers and across health systems to ensure the availability of a comprehensive pool of patients’ risk information and facilitate prompt clinical decision-making. Finally, collecting and integrating accurate data into the EHR system is key for developing and implementing an effective automated PrEP prescribing tool.

## Data Availability

Data cannot be shared publicly to protect participant's privacy. Data are available from the Institutional Review Board of the University of Florida for researchers who meet the criteria for access to confidential data.

